# Investigating chromosomal radiosensitivity in inborn errors of immunity: insights from DNA repair disorders and beyond

**DOI:** 10.1101/2024.08.14.24311337

**Authors:** Elien Beyls, Evi Duthoo, Lynn Backers, Karlien Claes, RAPID clinicians, Marieke De Bruyne, Lore Pottie, Victoria Bordon, Carolien Bonroy, Simon J Tavernier, Kathleen BM Claes, Anne Vral, Ans Baeyens, Filomeen Haerynck

**Affiliations:** Primary Immunodeficiency Research Lab (PIRL), Department of Internal Medicine and Pediatrics, Ghent University, Ghent, Belgium; Center for Primary Immunodeficiency, Jeffrey Modell Diagnosis and Research Center, Ghent University Hospital, Ghent, Belgium; Department of Human Structure and Repair, Ghent University, Ghent, Belgium; Center for Medical Genetics, Ghent University Hospital, Ghent, Belgium; Department of Biomolecular Medicine, Ghent University, Ghent, Belgium; Cancer Research Institute Ghent (CRIG), Ghent University Hospital, Ghent, Belgium; Oncology and Stem Cell Transplantation expert Center, Department of Pediatric Hematology, Ghent University Hospital, Ghent, Belgium; Department of Diagnostic Sciences, Ghent University, Ghent, Belgium; Department of Laboratory Medicine, Ghent University Hospital, Ghent, Belgium; Department of Biomedical Molecular Biology, Ghent University, Ghent, Belgium; Unit of Molecular Signal Transduction in Inflammation, VIB-UGent Center for Inflammation Research, Ghent, Belgium; Radiobiology Lab, Department of Human Structure and Repair, Ghent University, Ghent, Belgium; Department of Pediatric Respiratory and Infectious Medicine, Ghent University Hospital, Ghent, Belgium

**Keywords:** Radiosensitivity, human inborn errors of immunity (IEI), micronucleus assay, DNA repair

## Abstract

Human inborn errors of immunity (IEI) represent a diverse group of genetic disorders affecting the innate and/or adaptive immune system. Some IEI entities comprise defects in DNA repair factors, resulting in (severe) combined immunodeficiencies, bone marrow failure, predisposition to malignancies, and potentially result in radiosensitivity (RS). While other IEI subcategories such as common variable immunodeficiency (CVID) and immune dysregulation disorders also associate with lymphoproliferative and malignant complications, the occurrence of RS phenotypes in the broader IEI population is not well characterized. Nonetheless, identifying RS in IEI patients through functional testing is crucial to reconsider radiation-related therapeutic protocols and to improve overall patient management. This study aimed to investigate chromosomal RS in a diverse cohort of 107 IEI patients using the G0 cytokinesis-block micronucleus (MN) assay. Our findings indicate significant variability in RS across specific genetic and phenotypical subgroups. Severe RS was detected in all ataxia-telangiectasia (AT) patients, a FANCI deficient and ERCC6L2 deficient patient, but not in any other IEI patient included in this cohort. Age emerged as the single influencing factor for both spontaneous and radiation-induced MN yields, while the manifestation of additional clinical features, including infection susceptibility, immune dysregulation, or malignancies did not associate with increased MN levels. Our extensive analysis of RS in the IEI population underscores the clinical importance of RS assessment in AT patients and supports RS testing in all IEI patients suspected of having a DNA repair disorder associated with radiosensitivity.

## INTRODUCTION

Human inborn errors of immunity (IEI) encompass a diverse group of 485 disease-causing gene defects which are categorized into 10 subgroups (I-X) according to the International Union of Immunological Societies (IUIS). These subgroups comprise a range of overlapping clinical and immunological features including increased susceptibility to infections, autoimmunity, allergy, autoinflammatory diseases, and bone marrow failure. Additionally, IEIs often associate with lymphoproliferative disorders and early incidence of malignancy, primarily of lymphoid origin [1–4]. Despite advancements in diagnostic algorithms and next generation sequencing [5–7], the molecular diagnostic rate remains low due to individual rarity and heterogeneity of IEIs [8,9]. Nonetheless, early diagnosis is crucial for appropriate treatment and management, preventing life-threatening complications, and improving patient outcome [10].

For some IEI entities, the immunodeficiency is caused by a genetic defect in one of the DNA damage response (DDR) factors. While DNA double strand break (DSB) repair pathways are crucial for preserving genomic integrity against exogenous damage like ionizing radiation (IR), similar mechanisms are also utilized under normal physiological conditions. Development and diversification of the adaptive immune system relies on V(D)J recombination and class switch recombination (CSR), two somatic processes that require DSB repair. Depending on the DSB repair factors involved, IEI patients display a variable degree of clinical features, including a syndromic appearance, increased cancer predisposition, neurological deficits, and radiosensitivity (RS) [11–13]. Deficiencies in core components of the non-homologous end-joining (NHEJ) DSB repair pathway, such as Artemis and DNA ligase IV, are linked to severe combined immunodeficiency (SCID, group I) and RS [14,15]. The most radiosensitive syndromes result from defects in ataxia-telangiectasia mutated (*ATM*) and Nijmegen breakage syndrome 1 (*NBS1*) (categorized as syndromic combined immunodeficiencies (CID), group II) and both encode factors implicated in the initiation and coordination of DSB signaling, as well as homologous recombination (HR) repair [16].

Therapeutic and monitoring procedures, including diagnostic imaging, often involve the use of radiation or other genotoxic agents. However, radiosensitive IEI patients face an increased risk for adverse reactions towards conventional radiotherapy and conditioning regimens for hematopoietic stem cell transplantation (HSCT). Adapted therapeutic protocols are widely recognized for their ability to limit these severe radiation-induced toxicities and enhance overall patient prognosis [17–19]. Assessing the RS status of patients with a suspected DNA repair disorder through functional testing is thus highly recommended to guide the deliberate use of DSB-inducing agents [17,20]. Moreover, even with a definitive molecular diagnosis, functional RS testing remains important in DNA repair disorders [20]. Prediction of the radiation response based solely on the affected DNA repair gene is still challenging as mildly impacting variants (e.g. leaky SCID, variant AT) may result in variable expression of clinical features, including the RS phenotype [21–24]. Moreover, progressive implementation of genetic testing continuously uncovers variants in novel IEI-causing genes. In these diseases, RS may not have been recognized as a phenotypic component [25–28].

Interestingly, a few reports have described increased *in vitro* RS in IEI disorders beyond the well-known DSB repair syndromes. RS was suggested as a potential disease characteristic in ARPC1B deficiency, LRBA deficiency, and hyper-IgM syndromes, including AID and CD40LG deficiency [29–32]. An increase in RS was also reported in patients with genetically undefined common variable immunodeficiency (CVID, group III), although the underlying mechanism remains unknown [33–36]. Of note, an excess risk of lymphoproliferation and malignant diseases was documented for these phenotypic subgroups.

This study aimed to comprehensively analyze chromosomal RS across a diverse spectrum of IEI patients using the standard G0 cytokinesis-block micronucleus (MN) assay, which primarily evaluates NHEJ-dependent DSB repair. In addition to inclusion of patients with genetically confirmed DNA repair defects, we focused on the following specific diagnostic categories: SCID, syndromic CID, CVID with lymphoproliferative or malignant disease, and bone marrow failure (BMF). Here, we present the largest study to date exploring RS testing in IEIs beyond DNA repair syndromes, validating its clinical importance in diagnosing IEI patients.

## MATERIALS & METHODS

### Study approval

This study was reviewed and approved by the Ethics Committee of the Ghent University Hospital (reference no. 2012/593, 2019/0461, and 2019/1565). Written informed consent was obtained from all participants in this study, in accordance with the 1975 Helsinki Declaration.

### Study design and patient population

From January 2018 till March 2024, chromosomal RS was assessed in a cohort of patients with IEI at the Radiobiology Lab, Department of Human Structure and Repair, Ghent University (Belgium). RS testing was initially performed on IEI patients from Ghent University Hospital and was subsequently expanded to include patients from other Belgian hospitals. Both pediatric and adult (≥ 18 years) patients were included, identified with either a genetically or clinically confirmed IEI. Criteria provided by the European Society for Immunodeficiency (ESID) were used to establish a definite IEI diagnosis [37]. IEI patients were categorized into 10 subgroups (I-X) based on the IUIS phenotypical classification [2]. Patients were deemed not eligible for inclusion in the following cases: SCID patients with a maternal T cell engraftment and patients for whom an insufficient yield of binucleated (BN) cells were obtained upon MN scoring. Patients for whom an IUIS classification could not be determined due to insufficient available clinical information were also excluded. We additionally included healthy relatives carrying a heterozygous (likely) pathogenic variant in a known DNA repair gene.

Retrospective collection of demographic and clinical information for all patients included age at blood sampling for RS analysis, sex, age at onset of IEI symptoms, infection susceptibility, autoimmunity (autoimmune cytopenias, organ specific and systemic autoimmune diseases), and benign lymphoproliferation (overactivation of lymphoid organs: lymphadenopathies, splenomegaly, and hepatomegaly) as immune dysregulation phenotypes, consanguinity, and history of malignancy. During the time course of the study, information on the occurrence of malignancies or HSCT therapy post-RS testing was also collected. Information on concurrent therapies (e.g. immunoglobulin replacement therapy or immunosuppressive treatment) at the time of blood sampling was not recorded.

### Genetic analysis

EDTA blood was drawn from the patients (and relatives) for DNA extraction and downstream molecular studies using the MagCore® Genomic DNA Large Volume Whole Blood Kit (RBC Bioscience, Code 104) according to the manufacturer’s protocols. For the generation of whole exome sequencing (WES) data, we applied the SureSelectXT Human All Exon V7 (Agilent Technologies) or KAPA HyperExome V1 (Roche) kits for target enrichment. Sequencing was performed on HiSeq 3000 or Novaseq 6000 (Illumina).

The BWA-MEM 0.7.17 algorithm was used for read mapping against the human genome reference sequence (NCBI, GRCh38/hg38), duplicate read removal, and variant calling. Variant calling and filtering were performed using our in house developed analysis platform Seqplorer, a graphical web interface that executes SQLite Gemini queries on an underlying database through straightforward dropdown menus and presents the results in a clear manner (https://github.ugent.be/cmgg/seqplorer). The position of the called variants was based on NCBI build GRCh38. Nucleotide numbering was according to the Human Genome Variation Society guidelines (HGVS). Potential copy number variants (CNV) of exons or entire genes were called using ExomeDepth, an algorithm which uses exome data to detect read depth differences, and an in-house developed script [38]. Variants are classified using an in house developed tool based on the ACMG [39] and ACGS [40] guidelines in the following classes: class 1 = benign, class 2 = likely benign (>95% certainty that variant is benign), class 3 = variant of unknown significance, class 4 = likely pathogenic (>90% certainty that variant is pathogenic) and class 5 = pathogenic.

### Radiosensitivity assessment by the G0 cytokinesis-block micronucleus assay

A standard protocol for the G0 MN assay was used, as described previously (for a detailed protocol: see Supplementary Materials and Fig. S1) [41]. A group of 50 adult healthy individuals (38% male, median age 32 years, age range of 22-58 years) was recruited to constitute reference values for RS classification of IEI patients. In this study, we defined the criterium for chromosomal instability as spontaneous MN yields (0 Gy) exceeding the mean + 3SD of the reference healthy control group. To determine radiation-induced MN values, spontaneous MN yields were subtracted from the MN values after 0.5 Gy and 1 Gy irradiation. RS classification was based on the results of the 1 Gy radiation dose. Patients for whom the 1 Gy radiation-induced MN yields exceeded the mean + 3SD threshold of the reference healthy control group were considered as ‘severe radiosensitive’. ‘Intermediate radiosensitivity’ was indicated when these MN yields ranged between the mean +2SD and +3SD threshold. Patients with MN yields below the mean + 2SD threshold were designated as ‘not radiosensitive’. RS classification was based on the first sampling in all cases, although for some patients a second sample was obtained to investigate the reproducibility of the assay.

### Statistical analysis

Data were analyzed using GraphPad Prism Software (version 10). Spearman’s r was used to test correlation of variables. Associations of clinical parameters with differential MN yields were assessed by excluding patients with a confirmed defect in one of the DNA DSB repair-related genes. A two-tailed t-test or a Mann-Whitney test was used for these analyses, in accordance with the normality assumption. Statistical significance was set at p<0.05.

## RESULTS

*Personally identifiable patient information was redacted in accordance with medRxiv requirements*.

### Study population characteristics

IEI patients across all different IUIS subgroups were included, except autoinflammatory disorders (group VII) and complement deficiencies (group VIII). Predominantly antibody deficiencies (PAD) (IUIS group III) (n = 30, 28%) was the most common IUIS category (Table 1 and Fig. 1). Twenty-three patients (21%) were classified as CID with associated or syndromic features (group II), and 24 (22%) as BMF (group IX). The global male-to-female ratio was 1.02, with a median age of 15 years (range: 4 months to 61 years). Out of 107 patients, 69 (64%) were younger than 18 years at the time of RS testing. Consanguinity in the parents was recorded for a minority of the patients (n = 8, 7%). Most patients displayed an early childhood-onset of IEI symptoms: 21 (20%) before the age of one and 57 (53%) between 1-10 years of age. First symptoms started after 20 years of age for 27 patients (25%). Recurrent infections were common (n = 72, 67%) in all IUIS categories, except group IX. Notably, signs of immune dysregulation were noted in 42 patients (39%), categorized in groups I, II, III, IV, VI, and IX. Thirty-three (31%) and 18 (17%) patients suffered from autoimmunity and lymphoproliferation, respectively. Seventeen (16%) patients underwent hematopoietic stem cell transplantation (HSCT) post-RS testing, while a history of malignancy was recorded for 21 (20%) patients. The malignant neoplasms were primarily of hematological origin and included nephroblastoma (n = 1), Hodgkin lymphoma (n = 4), non-Hodgkin lymphoma (n = 4), leukemia (n = 3), and myelodysplastic syndrome (n = 9) (Table S1). Further details of the cohort are provided in Table 1 and Fig. 1.

**Figure 1.**
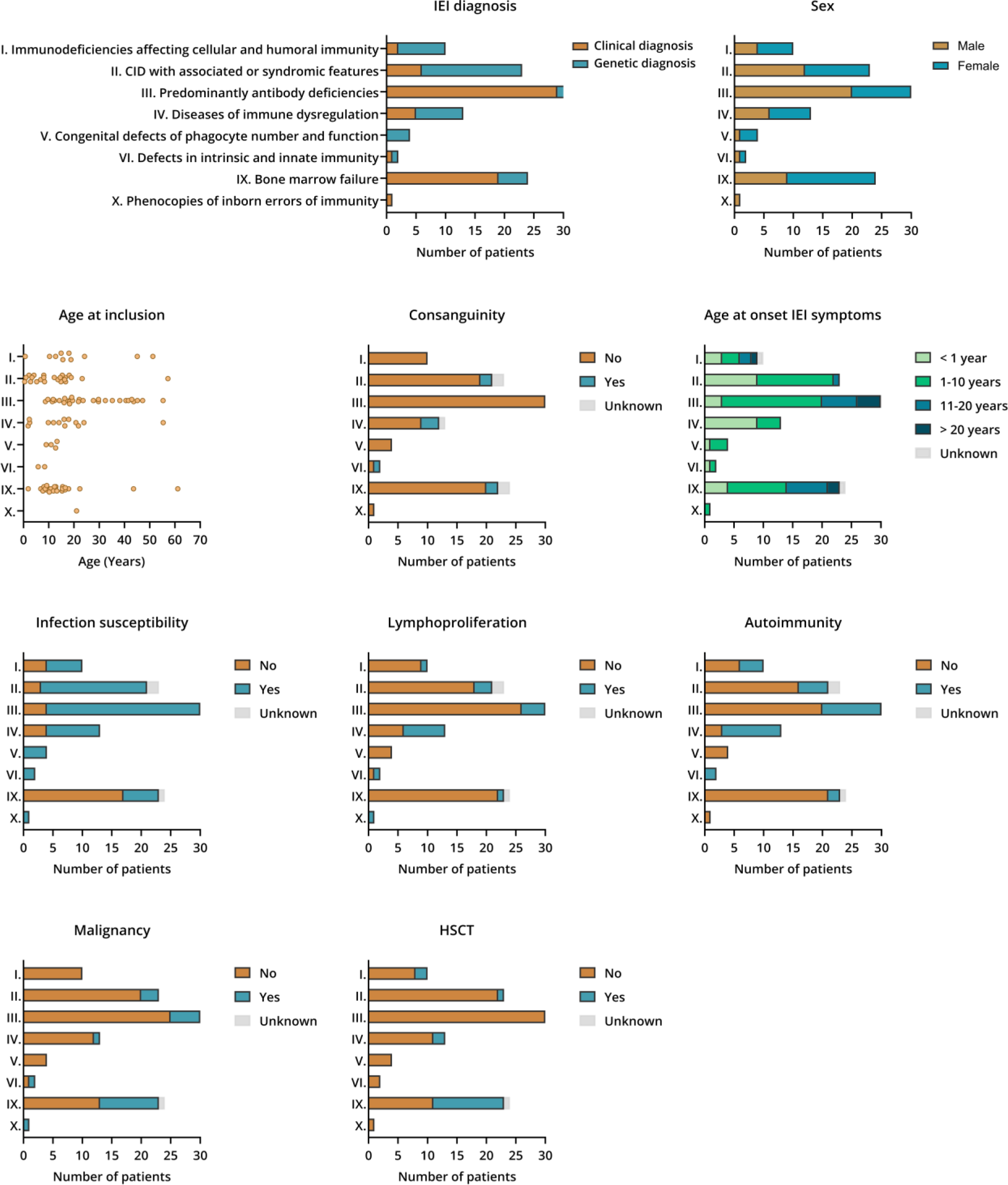
Study population characteristics of the inborn error of immunity (IEI) patient cohort (n = 107). Patients were classified according to the 10 IUIS subgroups of IEIs. Distribution of the patients across these subgroups is displayed, including the occurrence of the following clinical parameters and immunological manifestations: genetic or clinical IEI diagnosis, sex, age at inclusion, consanguinity, age at onset IEI symptoms, infection susceptibility, lymphoproliferation, autoimmunity, history of malignancy, and hematopoietic stem cell transplantation (HSCT).

**Table 1.**
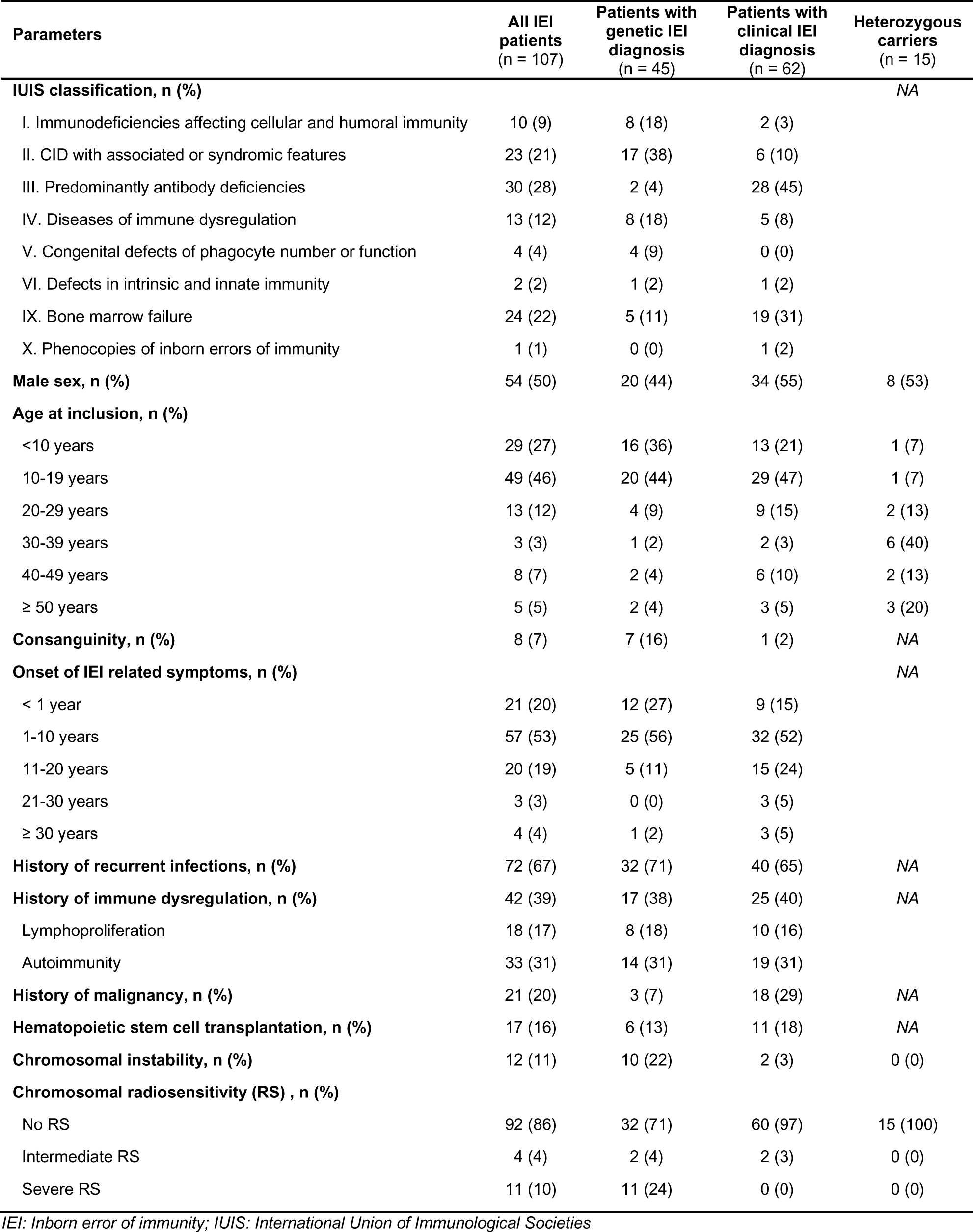
Demographic and clinical characteristics of IEI patients and heterozygous carriers.

| Parameters | All IEI patients<br>(n = 107) | Patients with<br>genetic IEI<br>diagnosis<br>(n = 45) | Patients with<br>clinical IEI<br>diagnosis<br>(n = 62) | Heterozygous<br>carriers<br>(n = 15) |
| --- | --- | --- | --- | --- |
| **IUIS classification, n (%)** |  |  |  | NA |
| I. Immunodeficiencies affecting cellular and humoral immunity | 10 (9) | 8 (18) | 2 (3) |  |
| II. CID with associated or syndromic features | 23 (21) | 17 (38) | 6 (10) |  |
| III. Predominantly antibody deficiencies | 30 (28) | 2 (4) | 28 (45) |  |
| IV. Diseases of immune dysregulation | 13 (12) | 8 (18) | 5 (8) |  |
| V. Congenital defects of phagocyte number or function | 4 (4) | 4 (9) | 0 (0) |  |
| VI. Defects in intrinsic and innate immunity | 2 (2) | 1 (2) | 1 (2) |  |
| IX. Bone marrow failure | 24 (22) | 5 (11) | 19 (31) |  |
| X. Phenocopies of inborn errors of immunity | 1 (1) | 0 (0) | 1 (2) |  |
| **Male sex, n (%)** | 54 (50) | 20 (44) | 34 (55) | 8 (53) |
| **Age at inclusion, n (%)** |  |  |  |  |
| <10 years | 29 (27) | 16 (36) | 13 (21) | 1 (7) |
| 10-19 years | 49 (46) | 20 (44) | 29 (47) | 1 (7) |
| 20-29 years | 13 (12) | 4 (9) | 9 (15) | 2 (13) |
| 30-39 years | 3 (3) | 1 (2) | 2 (3) | 6 (40) |
| 40-49 years | 8 (7) | 2 (4) | 6 (10) | 2 (13) |
| ≥ 50 years | 5 (5) | 2 (4) | 3 (5) | 3 (20) |
| **Consanguinity, n (%)** | 8 (7) | 7 (16) | 1 (2) | NA |
| **Onset of IEI related symptoms, n (%)** |  |  |  | NA |
| < 1 year | 21 (20) | 12 (27) | 9 (15) |  |
| 1-10 years | 57 (53) | 25 (56) | 32 (52) |  |
| 11-20 years | 20 (19) | 5 (11) | 15 (24) |  |
| 21-30 years | 3 (3) | 0 (0) | 3 (5) |  |
| ≥ 30 years | 4 (4) | 1 (2) | 3 (5) |  |
| **History of recurrent infections, n (%)** | 72 (67) | 32 (71) | 40 (65) | NA |
| **History of immune dysregulation, n (%)** | 42 (39) | 17 (38) | 25 (40) | NA |
| Lymphoproliferation | 18 (17) | 8 (18) | 10 (16) |  |
| Autoimmunity | 33 (31) | 14 (31) | 19 (31) |  |
| **History of malignancy, n (%)** | 21 (20) | 3 (7) | 18 (29) | NA |
| **Hematopoietic stem cell transplantation, n (%)** | 17 (16) | 6 (13) | 11 (18) | NA |
| **Chromosomal instability, n (%)** | 12 (11) | 10 (22) | 2 (3) | 0 (0) |
| **Chromosomal radiosensitivity (RS) , n (%)** |  |  |  |  |
| No RS | 92 (86) | 32 (71) | 60 (97) | 15 (100) |
| Intermediate RS | 4 (4) | 2 (4) | 2 (3) | 0 (0) |
| Severe RS | 11 (10) | 11 (24) | 0 (0) | 0 (0) |

IEI: Inborn error of immunity; IUIS: International Union of Immunological Societies

A definitive genetic diagnosis was obtained for 45 patients (42%), either prior to RS testing or during follow-up. The diagnostic yield was particularly low in groups III and IX (Table 1 and Fig. 1). We identified 52 pathogenic variants across 21 distinct IEI-related genes, including 18 novel variants (not reported in ClinVar). Five out of these 21 genes are well-described to be implicated within one of the DNA DSB repair pathways [20], affecting a total of 18 patients. Within group II, 9 patients harbored biallelic defects in *ATM* and two harbored biallelic alterations in *BLM* [42]. Biallelic inactivation was detected in 5 patients of group IX (BMF): *FANCA* (n = 3), *FANCI* (n = 1), or *ERCC6L2* (n = 1). Several other IEI-causing variants were identified more than once: *LRBA* (n = 5), *IKZF1* (n = 6) [43], *SBDS* (n = 3), *RNU4ATAC* (n = 2) [44], and *STAT3* (gain-of-function, GOF) (n = 2). The remaining 11 genetic disorders were found in single cases, involving pathogenic variants in *IRAK4*, *NFKB1*, *PIK3CD* (GOF), *USB1*, *TTC37*, *UNC13D*, *SLC46A1*, *ADA*, *RMRP*, *WAS* (loss-of-function, LOF), and *CD40LG*. Full details regarding all identified variants are provided in Tables S2-8. Additionally, RS analysis was performed for 12 healthy heterozygous *ATM* carriers (Table S9), and three healthy heterozygous relatives of an Artemis patient (*DCLRE1C*), previously described by Strubbe et al. [45].

### Evaluation of chromosomal instability and radiosensitivity within the total IEI cohort

Spontaneous and radiation-induced MN yields for 1 Gy are presented for the entire healthy control and IEI patient population, together with the threshold MN values for chromosomal instability and chromosomal RS (Fig. 2a). Next to IUIS group I (immunodeficiencies affecting cellular and humoral immunity), groups II (CID with syndromic features) and IX (BMF) are well-known to encompass monogenetic disorders of the DNA DSB repair pathways [3]. Among the 12 patients with elevated spontaneous MN values, one was categorized in group I, 9 in group II, and two in group III (PAD). Analysis of radiation-induced MN scores showed that chromosomal RS was absent in the majority of the patients (n = 92, 86%) (Fig. 2a). Intermediate RS was found in 4 patients: one patient in groups I, II, IV (diseases of immune dysregulation), and IX. Eleven patients were considered as severe radiosensitive, of which 9 were categorized in group II and two in group IX. Importantly, none of the healthy controls exceeded the mean + 3SD threshold for 0 Gy or the mean + 2SD threshold for both radiation doses (Fig. 2a and Fig. S2a). For all doses, the coefficients of variation (CV) of the reference MN values were in accordance with those previously reported for healthy individuals (Table 2) [46,47]. When considering all healthy controls and patients together, a moderate but significant correlation (r = 0.60, p<0.0001) was observed between the radiation-induced MN yields of 0.5 Gy and 1 Gy (Fig. S2b). Given that the comparison between the two radiation doses did not reveal major discrepancies, a single dose (1 Gy) can be effectively used for RS classification.

**Figure 2.**
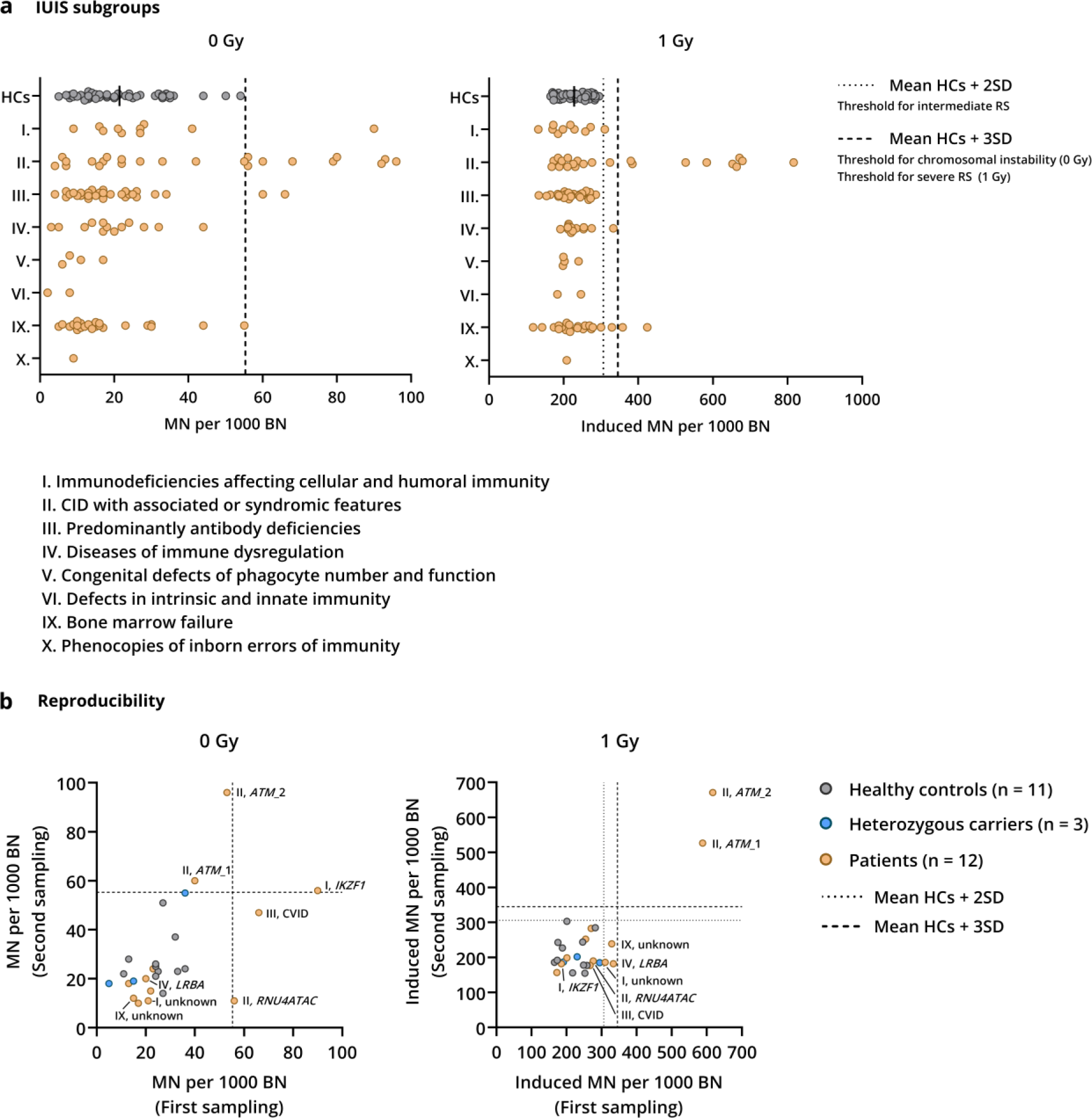
Evaluation of chromosomal instability and radiosensitivity (RS) of the entire IEI patient cohort. **(a)** Spontaneous (0 Gy) and radiation-induced (1 Gy) MN yields are displayed per IUIS subgroup for all patients. Chromosomal instability was indicated when MN values exceed the mean + 3SD threshold (dashed line) for 0 Gy. Patients with radiation-induced MN yields (1 Gy) higher than the mean + 2SD threshold (dotted line) or higher than the mean + 3SD threshold (dashed line) were classified as intermediate or severe radiosensitive, respectively. Patients were considered as not radiosensitive when the MN yields were lower than the mean + 2SD threshold. **(b)** Reproducibility of the G0 MN assay was assessed by comparing spontaneous (0 Gy) and radiation-induced (1 Gy) MN yields of the first and second sampling for 11 healthy controls, 3 heterozygous carriers, and 12 patients. Patients that exceeded the 0 Gy and/or 1 Gy threshold are annotated on both panels.

**Table 2.**
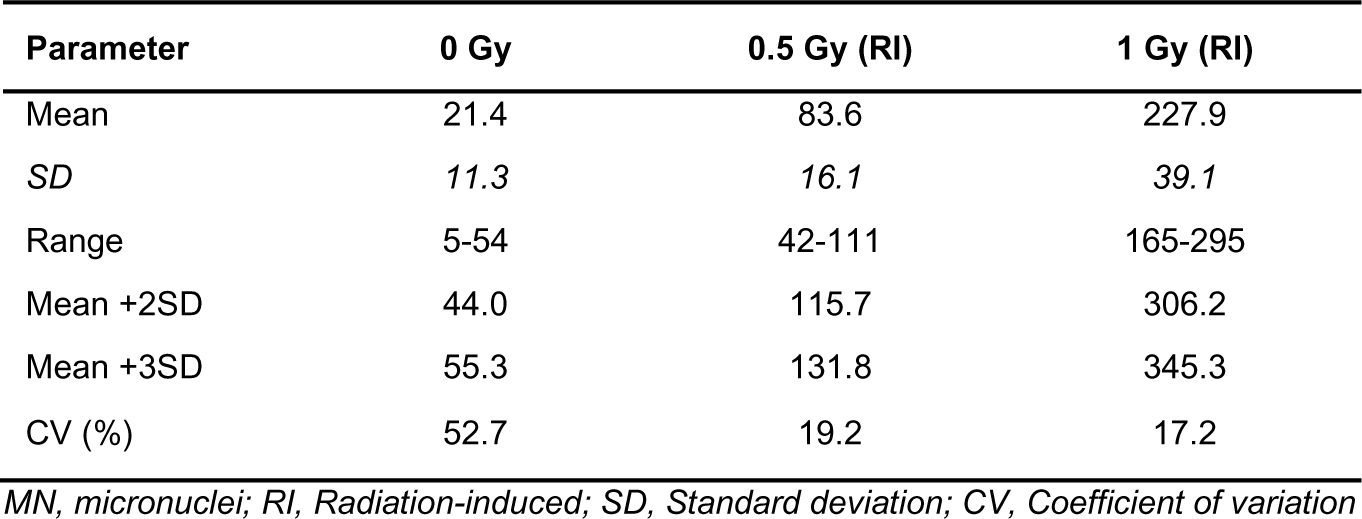
Reference G0 MN values of the healthy control population (n = 50).

To investigate the reproducibility of the categorization of chromosomal instability and RS used in this study, a second sample was analyzed for 11 healthy controls, 3 heterozygous carriers, and 12 patients (Fig 2b and Fig. S2c). For all examined healthy controls (n = 11) and heterozygous mutation carriers (n = 3), repeated sampling confirmed the absence of chromosomal instability or RS phenotypes (Fig. 2b). Of the 12 retested IEI patients, interpretation of chromosomal instability was inconsistent for 4 patients as spontaneous MN yields exceeded the threshold in only one of the two samples. Importantly, for two of the eleven severe RS patients, a second sampling readily confirmed this initial classification (Fig 2b). Of the 4 patients initially identified as intermediate radiosensitive, we were able to retest three patients, revealing a non-RS phenotype for the second sample (Fig. 2b). It must be noted that the criterium for intermediate RS is strict due to the narrow range between the mean +2SD and +3SD threshold (306-345 MN per 1000 BN). The results of the repeated sampling prompted us to interpret the intermediate RS category with more caution. Taken together, these findings demonstrate the reliability of the G0 MN assay to identify severe RS phenotypes but suggest that a second sampling is required for cases with intermediate RS results.

### IEI patients with specific DNA repair gene defects display chromosomal instability and radiosensitivity

Within group I, increased spontaneous MN values were detected in one of the 6 related patients with an Ikaros deficiency (*IKZF1*) (Fig. 2b and Fig. 3a). The absence of a chromosomal instability phenotype in the affected relatives of this patient (female; 40-49 years) suggests that this increase does not originate from the *IKZF1* variant. Intermediate RS was observed in another patient presenting with a CID phenotype of unknown cause (group I) (Fig. 2b, Fig. 3a, and Fig. S3a). Due to HSCT therapy early in life, we were unable to perform lymphocyte-based RS analysis in patients with defects in core NHEJ factors causing T^−^B^−^ RS-SCID, such as Artemis [3].

**Figure 3.**
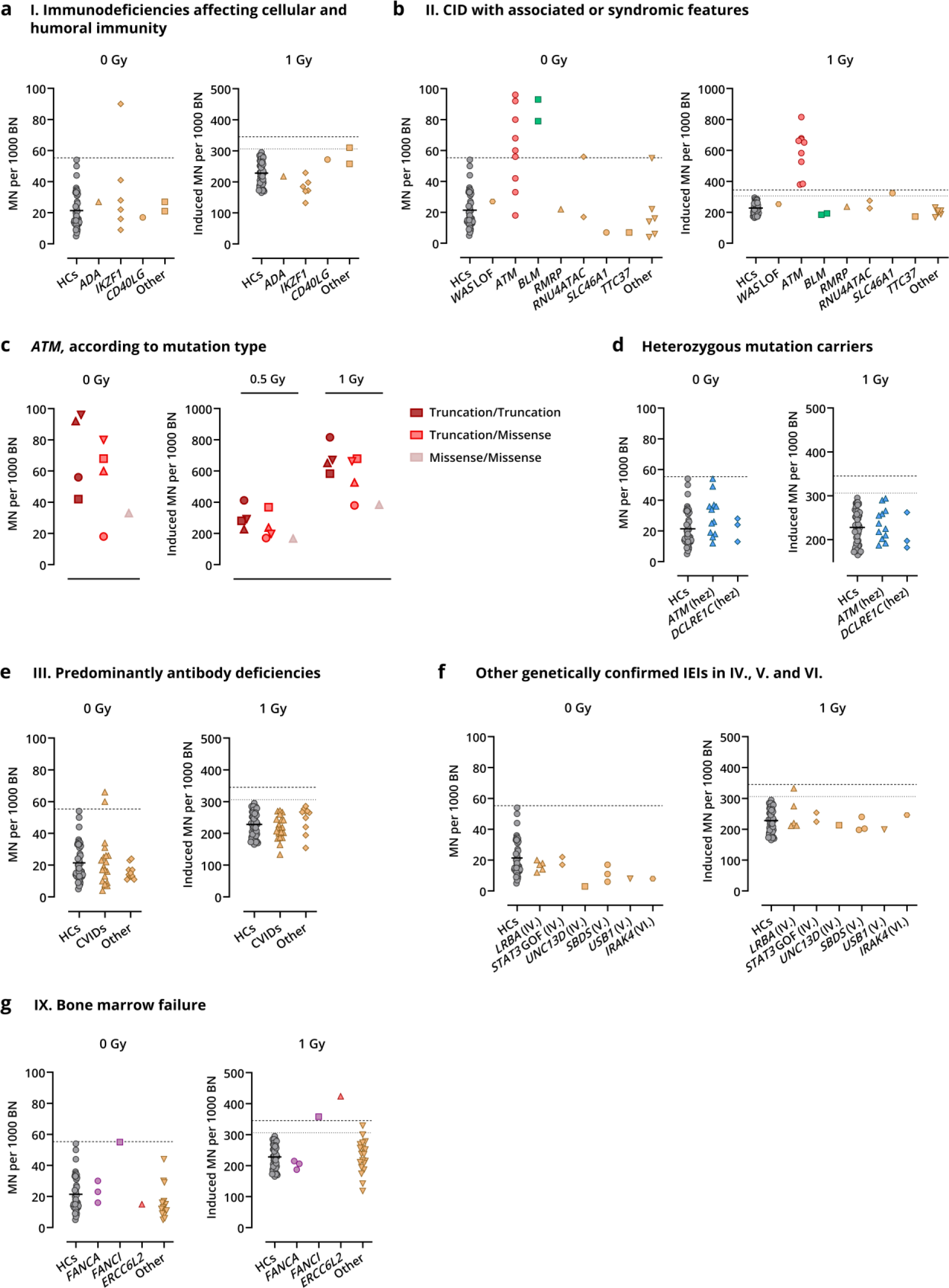
Chromosomal instability and radiosensitivity phenotypes according to specific gene defects. Spontaneous (0 Gy) and radiation-induced MN yields (1 Gy) are shown for healthy controls (HCs), heterozygous mutation carriers and patients with a confirmed genetic diagnosis, displayed per IUIS group. **(a)** Patients with a confirmed genetic defect in IUIS group I (immunodeficiencies affecting cellular and humoral immunity) **(b)** Patients with an identified defect in IUIS group II (combined immunodeficiency (CID) with associated or syndromic features). **(c)** Variability in MN results (0 Gy, 0.5 Gy, and 1 Gy) among the 9 ataxia telangiectasia (AT) patients, distinguished according to the mutation type. Four patients carried two truncating variants, one harbored a biallelic missense variant, and 4 were compound heterozygous for a truncating and a non-truncating variant. **(d)** Relatives of IEI patients carrying a heterozygous mutation in *ATM* or *DCLRE1C* (Artemis). **(e)** Patients classified in group III (predominantly antibody deficiencies), displayed separately for common variable immunodeficiency (CVID) and non-CVID (other) patients. **(f)** Patients with a confirmed genetic defect in IUIS group IV (diseases of immune dysregulation), group V (congenital defects in phagocyte number and function) or group VI (defects in intrinsic and innate immunity). **(g)** Patients with an identified genetic defect, categorized in IUIS group IX (bone marrow failure). Dotted and dashed lines indicate the mean + 2SD and mean + 3SD threshold values, respectively.

For both patients deficient in BLM (group II), a protein involved in HR-dependent DSB repair, a pronounced elevation in spontaneous MN was detected, albeit radiation-induced MN yields were not increased (Fig. 3b and Fig. S3b). All 9 radiosensitive cases within group II harbored biallelic *ATM* mutations. As AT is a known severe RS syndrome, these data support the validity of the mean + 3SD value as a threshold for RS. Importantly, the severe RS classification of one AT patient – identified with a novel homozygous *ATM* variant of uncertain significance (VUS) – underscores the added value of RS testing in supporting the pathogenic character of VUS (Table S3). Notably, considerable variability was noted among MN results of the AT patients (Fig 3c). Spontaneous MN values were only elevated for 6 of the 9 AT patients, with values varying from 18-96 MN per 1000 BN cells. Similarly, a broad range of radiation-induced MN scores was observed: 168-412 and 380-816 MN per 1000 BN for 0.5 and 1 Gy, respectively. These MN yields correspond to a fold increase over healthy controls ranging from 1.7-4.9, consistent with previous reports [48,49]. Despite this variability, the G0 MN assay displayed sufficient sensitivity to effectively categorize all AT patients as severe RS. A more detailed analysis of the MN data based on the variant type suggested a genotype-phenotype correlation. Patients carrying two truncating *ATM* variants appeared to display higher MN scores for both spontaneous and radiation-induced conditions compared to a patient with biallelic missense variants, showing a more attenuated RS phenotype. MN scores of patients with both truncating and missense *ATM* variants in a compound heterozygous state ranged between these extremes. These results suggest that the variant type in *ATM* contributed to the different levels of chromosomal RS.

Increased spontaneous MN values were detected in a patient with Roifman syndrome (*RNU4ATAC*, group II), but not in the patient’s sibling (Fig. 2b and Fig. 3b). Intriguingly, one patient with SLC46A1 deficiency (group II), causing hereditary folate malabsorption, displayed an intermediate RS phenotype, but we were unable to repeat the MN assay for this patient (Fig. 3b, and Fig. S3b). RS has not been studied in this specific disorder, although folate deficiency was described to interfere with DNA integrity by causing various types of DNA breaks that are not repaired efficiently [50,51].

No chromosomal instability or RS phenotypes were observed among the heterozygous carriers of a monoallelic variant in either *DCLRE1C* (n = 3) or *ATM* (n = 12) (Fig. 3d and Fig. S3c).

Within group III (PAD), twenty patients were diagnosed with CVID. While no RS phenotypes were detected, two CVID patients (age range: 40-49 years) with unidentified genetic defects displayed chromosomal instability (Fig. 2b, Fig. 3e and Fig. S3d). Neither patient displayed signs of immune dysregulation (autoimmunity and lymphoproliferation) or had a history of malignancy.

Apart from one of the 5 LRBA patients with an intermediate RS phenotype (group IV, diseases of immune dysregulation), no other increases in spontaneous or radiation-induced MN yields were observed in the patients categorized in group IV, V (congenital defects of phagocyte number and function), or VI (defects in intrinsic and innate immunity) (Fig. 2b, Fig. 3f and Fig. S3e).

All three Fanconi anemia (FA, group IX, BMF) patients with pathogenic variants in *FANCA* showed normal spontaneous and radiation-induced MN yields, in contrast to one *FANCI* patient who displayed a severe RS phenotype (Fig. 3g and Fig. S3f). This patient developed a secondary malignancy (acute myeloid leukemia) at age 44 and succumbed to infectious complications during induction chemotherapy. Due to the rarity of reported *FANCI* cases and the lack of *in vitro* data, it remains unclear whether these clinical events and the observed chromosomal RS are connected [52]. Of note, impaired interstrand crosslink (ICL) DNA repair is a known hallmark of FA, which can be demonstrated by testing sensitivity to the interstrand crosslinkers Mitomycin C (MMC) or diepoxybutane (DEB) [53]. All FA patients exhibited MMC sensitivity (data not shown), supporting a definite diagnosis of FA. Intriguingly, a second patient in group IX exhibited severe RS with normal spontaneous MN values (Fig. 3g and Fig. S3f). Five years post-RS analysis, this patient was identified with a pathogenic biallelic *ERCC6L2* variant. Unfortunately, HSCT prevented repeated analysis to confirm these results. One patient with BMF of unknown cause showed intermediately elevated radiation-induced MN values, but not upon re-analysis (Fig. 2b, Fig. 3g, and Fig. S3f).

### Absence of significant association between MN yields and clinical manifestations

Given the heterogeneity observed in immunological and clinical features among IEI patients, we conducted an extended analysis to explore potential associations between these features and differences in spontaneous or radiation induced MN yields (Fig. 4 and Fig. S4). Interestingly, no significant differences in MN yields were detected between male and female patients (Fig. 4a and Fig. S4a).

**Figure 4.**
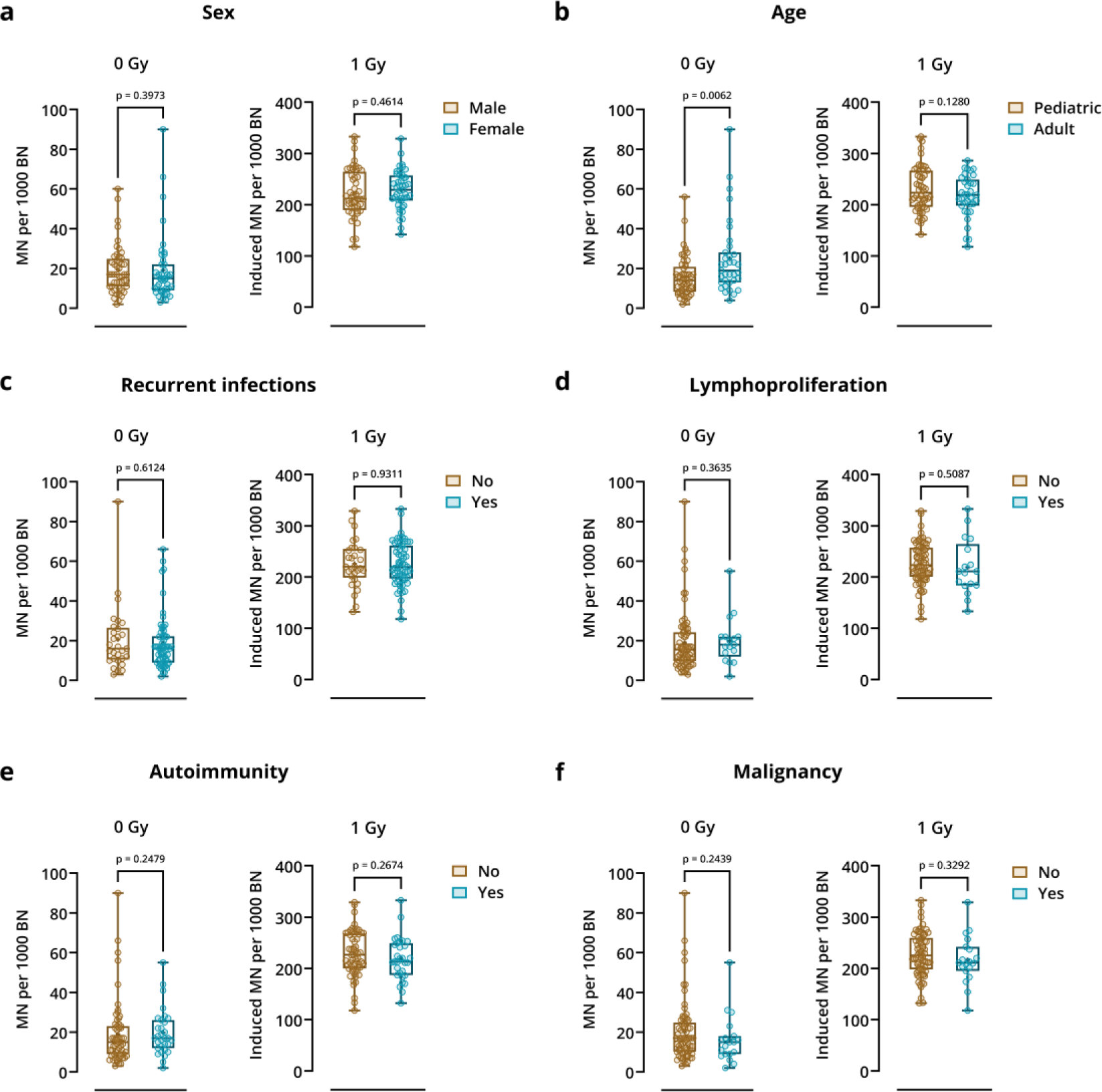
The occurrence of IEI-related co-morbidities are not associated with increased spontaneous or radiation-induced MN frequencies. Association between MN yields and the following clinical features and immunological manifestations were investigated: (**a**) sex, (**b**) age at inclusion, (**c**) recurrent infections, (**d**) lymphoproliferation, (**e**) autoimmunity, and (**f**) history of malignancy. Spontaneous (0 Gy) and radiation-induced MN yields (1 Gy) are displayed for each parameter. Boxplots depict the median, lower and upper quartiles of the MN yields, whiskers indicate minimum and maximum values. Statistical significance was assessed using the Mann-Whitney test (0 Gy) or unpaired t-tests (1 Gy). Patients with a confirmed defect in one of the DNA DSB repair-related genes were excluded for this analysis.

Pediatric patients (<18 years at time of RS analysis) exhibited lower levels of spontaneous MN, accompanied with higher levels of radiation-induced MN compared to adult patients (Fig. 4b and Fig. S4b). While MN yields did not appear to be affected by the occurrence of recurrent infections and lymphoproliferative disorders, patients with autoimmunity unexpectedly exhibited lower MN values compared to patients without autoimmune conditions, an observation that was only significant for the 0.5 Gy irradiated samples (Fig. 4c-e and Fig. S4c-e). A history of malignancy in IEI patients, diagnosed either before or after RS testing, did not associate with a difference in MN scores (Fig. 4f and Fig. S4f).

### MN yields correlate with age

Considering that the comparison between pediatric and adult IEI patients suggested a potential correlation between age and MN yields, we performed a more in-depth analysis using a pooled dataset of patients, healthy controls, and heterozygous mutation carriers (Fig. 5 and Fig. S5). Consistent with previous reports, we observed that spontaneous MN yields significantly increased with age (Fig. 5a) [54–57]. Markedly, 0.5 and 1 Gy radiation-induced MN yields tended to decrease with age, showing a weak negative correlation for both radiation doses. This correlation was also observed when considering all individuals together (healthy controls, patients, and heterozygotes) (Fig. 5b and Fig. S5). Although careful interpretation is warranted due to the clinical diversity of the individuals in our dataset, these results indicate that pediatric IEI patients are potentially characterized by a slightly increased chromosomal RS profile compared to adult IEI patients and adult healthy controls.

**Figure 5.**
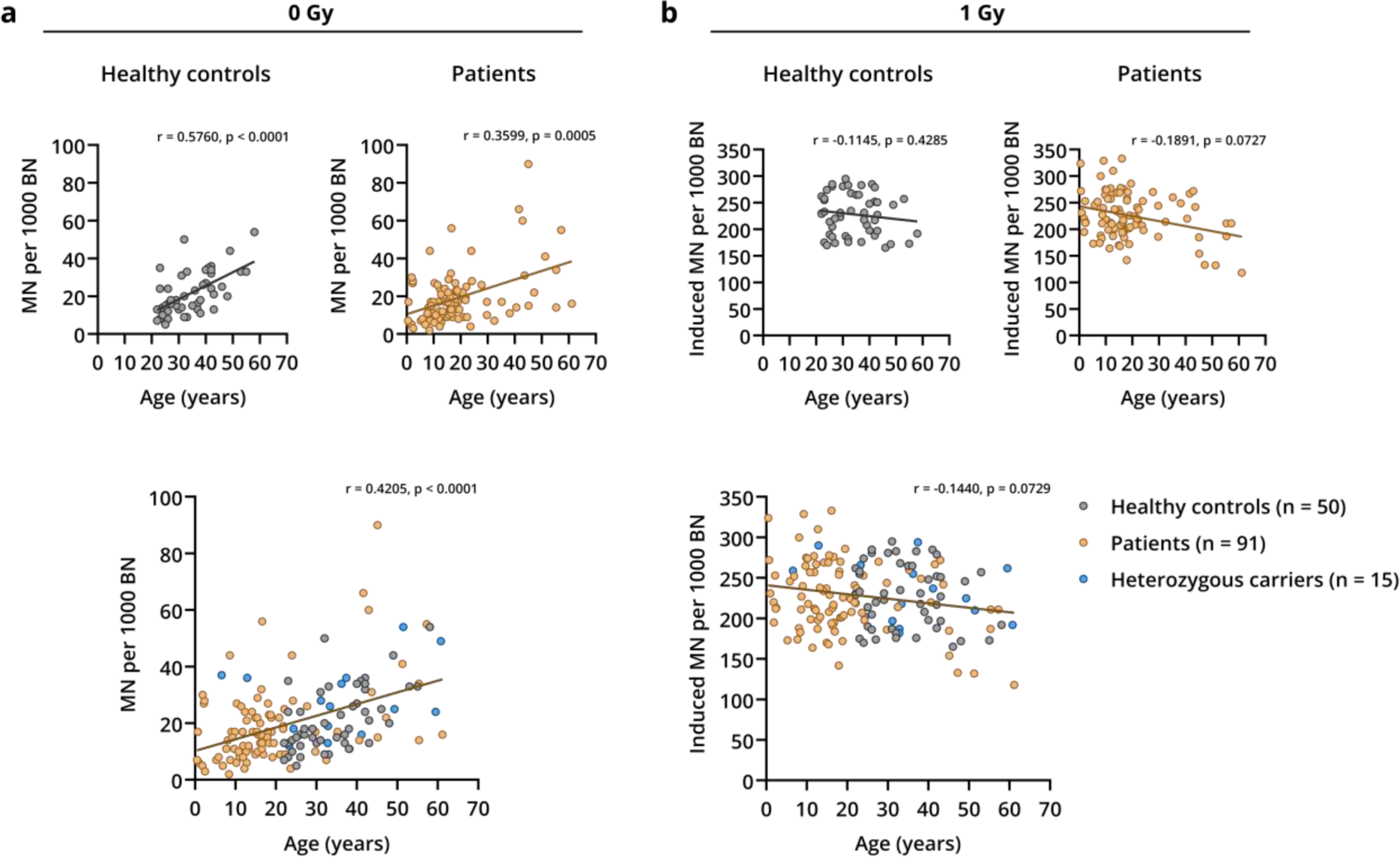
Age-related variation in spontaneous and radiation-induced MN yields in the entire cohort of IEI patients, heterozygous carriers, and healthy controls. A correlation analysis was performed between age and **(a)** spontaneous (0 Gy) and **(b)** radiation-induced MN yields (1 Gy), by calculating the Spearman’s rank correlation coefficient (r). The healthy control and IEI patient cohort were analyzed separately, as well as the entire study population together (healthy controls, patients, and heterozygous carriers). Patients with a confirmed defect in one of the DNA DSB repair-related genes were excluded for this analysis.

## DISCUSSION

Among the 10 phenotypic categories in the IUIS classification for IEIs, three subgroups include defects in DSB repair pathways [3]. Although radiosensitivity (RS) is often considered a characteristic of these subgroups – with ataxia telangiectasia (AT) as the prototypic RS disorder – the characterization and understanding of RS in the broader IEI population, including certain DNA repair disorders, remains limited. Here, we conducted a comprehensive investigation into chromosomal RS across a diverse range of IEIs, including patients with confirmed or suspected DSB repair defects and those with additional complications such as lymphoproliferation and/or malignant diseases. The principal findings of this study include: (1) a potential genotype-phenotype correlation among the severe RS AT patients as indicated by the striking variability in MN yields, (2) the detection of increased RS in single patients with a FANCI-deficiency and ERCC6L2-deficiency, (3) the lack of severe RS phenotypes among all the other IEI patients tested in this cohort, and (4) the observation that the investigated clinical and immunological features did not correlate with increased MN yields, except for age. These findings provide additional insights into the chromosomal RS status associated with specific DNA repair defects and underscore the need for guidelines on RS testing within the IEI patient population for routine clinical practice, to improve both diagnosis and management.

Cytogenetic tests such as the G0 MN assay have repeatedly been recognized for their potential to detect RS in AT patients [48,49,58,59]. In our analysis of patients with syndromic CID (group II), all AT patients included indeed displayed a distinct RS phenotype. Our data additionally suggested a correlation between MN yields and the underlying *ATM* mutation type, indicating a potential link between the genotype and the degree of chromosomal RS. Analogous, the spectrum of clinical manifestations in AT patients correlates with *ATM* expression levels, with severe phenotypic patients generally harboring nonsense or truncating variants and milder or atypical cases harboring missense or splice-site variants, which often allow some kinase activity [60–62]. Presumably, a high variety in chromosomal RS among AT patients could reflect a clinically relevant range of radiation responses. Given that the G0 MN assay effectively differentiated between grades of severe RS and proved valuable in validating novel VUS, we recommend RS assessment as a crucial component in the diagnostic process of all newly identified AT patients.

Although *ATM* carriers are well-known to have a relatively higher risk for breast cancer, their association with increased *in vitro* RS remains unclear. While some studies, applying other RS assays, reported differentiation between *ATM* heterozygous carriers and healthy controls [48,63], a previous study using the G0 MN assay was not able to demonstrate increased RS, consistent with our findings [48]. Generally, *ATM* carriers do not exhibit an increase in clinical adverse responses to radiation and do not require adaptations in treatment protocols involving IR exposure [64].

Chromosomal instability, predisposition to early-onset cancer, and variable immunodeficiency are common features of Bloom syndrome (group II). The BLM protein has a well-defined role in the HR pathway for DSB repair, with the association of Bloom syndrome with RS remaining controversial in literature [25,65]. The two patients presented here showed a profound increase in spontaneous MN, but did not display RS. An increase in sister chromatid exchanges (SCEs) represents the standard diagnostic marker, although our results also support the diagnostic value of spontaneous MN for determining chromosomal instability in Bloom syndrome patients [20].

To date, 22 causal genes for Fanconi anemia (FA, group IX) have been identified. FA is associated with bone marrow failure (BMF), increased cancer predisposition, and a range of developmental abnormalities. Many FA proteins are involved in both the HR and ICL repair pathway [66]. Despite the description of FA as a chromosomal instability syndrome, we did not detect increased spontaneous MN values in any of the 4 FA patients. Interestingly, chromosomal RS was absent in all three *FANCA* patients, while one *FANCI* patient presented with severe RS. These results indicate that the G0 MN test does not emerge as a suitable diagnostic marker for this syndromes, in contrast to ICL sensitivity assays [53,66]. Due to the large number of genes involved and the variety of RS assays used, *in vitro* studies have been unable to consistently demonstrate RS in FA patient cells [25,52,67–69].

ERCC6L2 deficiency, an autosomal recessive inherited BMF syndrome, was first identified by Tummala et al. and is clinically characterized by developmental abnormalities, progression to myelodysplastic syndrome, and acute myeloid leukemia, often necessitating HSCT therapy [70]. Multifaceted roles were considered for ERCC6L2, including a contribution to NHEJ repair. Similar to the severe RS phenotype exhibited by our ERCC6L2 patient, Zhang et al. reported an increased sensitivity to IR in SV40-transformed ERCC6L2 deficient fibroblasts [21]. Further research into this disease is warranted as ERCC6L2 deficiency may underlie excessive radiation-related toxicities, which was recently described in a ERCC6L2 deficient breast cancer patient [71].

Besides *ATM, ERCC6L2, BLM, FANCA,* and *FANCI*, none of the other 16 IEI-causing genes identified in this study have defined roles in DSB repair. Accordingly, severe RS was not detected in any of the patients affected by these other IEIs. In single studies, RS was demonstrated for patients deficient for LRBA (6/11 patients), ADA (1/1 patient), WASP (1/1 patient), and CD40LG (8/11 patients) [30–32,69]. In contrast to those previous reports, we did not observe severe RS in any of the 5 LRBA patients, nor in the single patients deficient for ADA, WASP, or CD40LG. Of note, one LRBA patient showed intermediate RS based on the first sample, but not upon retesting. Moreover, our group of CVID patients did not exhibit differences in radiation-induced MN yields as compared to healthy controls, contrary to reports of significant increases in RS in four other CVID cohorts [33–36]. However, it should be noted that RS was tested using another methodology throughout most of these studies. As cells are irradiated in the G2 and G0 phase respectively, the G2 chromatid and G0 MN assay may produce distinct results due to evaluation of cell cycle-specific repair mechanisms and different cell cycle checkpoints. For instance, Schrank et al. described that the nuclear actin polymerization factors WASP and the ARP2/3 complex are specifically required for HR, but not NHEJ [72]. Accordingly, increased levels of G2 chromatid-type aberrations were recently reported in patients deficient for ARPC1B and WASP [30].

CIDs, CVIDs, bone marrow failures, and immune dysregulation disorders represent the main IEI subtypes predisposing to malignancies [19,73,74]. While genomic instability and defective DNA repair are recognized as key factors in the tumorigenesis of monogenetic diseases such as AT, Bloom syndrome, and FA, the genetic etiology and mechanisms driving oncogenic processes in other IEIs remain largely undefined [73,75]. We could not document a potential link between chromosomal instability or RS and the occurrence of lymphoproliferative disorders or malignancies in our IEI patient cohort. Consequently, our findings do not allow to further define whether genomic instability and/or disrupted DNA DSB repair are key cancer hallmarks for these IEI entities.

In line with our data, a progressive increase in spontaneous MN frequencies has been reported in ageing individuals [54–57]. However, few studies have explored the effects of ageing on *in vitro* RS or extended the investigation by including pediatric individuals, with some controversy remaining due to small cohorts, limited age ranges, or variable methodologies [76–79]. Although our study cohort mainly consisted of IEI patients, the availability of MN data of both children and adults offered a valuable opportunity to investigate the effects of ageing more thoroughly. We observed a minor but negative correlation between age and radiation-induced MN yields, suggesting higher *in vitro* sensitivity to IR in younger individuals. Interestingly, multiple reports identified ageing and sex as confounding factors, with sex affecting MN yields in older adults but not in children [55,57]. Consistent with this, we found no significant differences between male and female patients in our predominantly pediatric cohort. Together, these results point towards the requirement of age-matched, but not sex-matched, controls to obtain references values for analyzing chromosomal instability and/or RS.

Although the G0 MN assay is a well-established cytogenetic tool with extensive applications in radiobiological research, its clinical implementations can be limited due to intrinsic inter- and intra-individual variability [80–85]. In this study, severe RS phenotypes were reliably identified, but the substantial individual variability prevented consistent and unequivocal interpretation of the intermediate RS category. A second sampling is thus strongly recommended when intermediate RS phenotypes are encountered in the initial analysis. Nevertheless, MN form a valuable cellular endpoint for RS as they represent both unrepaired and misrepaired DSBs, indicative for radiation-induced cell death and mutagenic processes, respectively. Importantly, MN yields remain to be used with caution as general predictive biomarkers of radiation-related adverse tissue reactions and radiation-induced cancer risks [86].

Based on our extensive investigation into chromosomal RS within the IEI population, we recommend RS testing in individual cases with concerns of an underlying DNA repair defect, especially prior to the therapeutic use of IR. Although the associated phenotypic features of these patients are often heterogeneous, early manifestation of an immunodeficiency in addition to a syndromic appearance, developmental and neurologic abnormalities, malignant disease, and/or BMF should raise suspicion of such inherited disorder [20]. We additionally emphasize the need for special awareness towards highly rare or newly identified IEI-associated genes with unknown RS associations. Our study also supports the use of RS testing as a guiding tool for genetic analysis. RS assays can aid in the validation of pathogenic VUS in known RS-associated genes (e.g. *ATM*) or can be indicative for the involvement of a DNA repair defect (e.g. *ERCC6L2*).

### Statements

Personally identifiable patient information was redacted in accordance with medRxiv requirements. IDs used in this study were not known to anyone outside the research group.

## Acknowledgements

We gratefully thank all patients and family for consenting to this research. The authors would also like to thank L. Pieters, G. De Smet, T. Thiron, J. Aernoudt, and E. Bes for performing the laboratory work and MN scoring. This work is supported by TBM-FWO RAPID. The Center for Primary Immune deficiency is recognized as a European Reference Network for Rare Immunodeficiency, Autoinflammatory, and Autoimmune Diseases Network (ERN-RITA) and is additionally supported by the Jeffrey Modell Diagnostic and Research foundation.

## Author contributions

A.V., A.B., C.B., V.B., F.H., and K.B.M.C. acquired funding and conceptualized, planned, and supervised the study. L.B., M.B., L.P. and K.B.M.C. contributed to genetic data analysis. A.V. and A.B. supervised the radiosensitivity testing. E.B. and E.D. analyzed the radiosensitivity data, collected and analyzed clinical data, and drafted the manuscript and figures. V.B., F.H., and all RAPID clinicians included patients, provided patient material, and helped collecting clinical information. K.C. informed patients about the study and collected informed consent approved by the ethics committee. E.B., E.D., A.B., A.V., K.B.M.C., S.J.T., and F.H. provided scientific guidance and critical discussion. All authors contributed to critical review of the manuscript and approved the final version.

## Data availability

The data generated and/or analyzed in the present study is available upon reasonable request to the corresponding authors.

## DECLARATIONS

### Ethics approval

This study was reviewed and approved by the Ethics Committee of the Ghent University Hospital (reference no. 2012/593, 2019/0461 and 2019/1565). Written informed consent was obtained from all participants in this study, in accordance with the 1975 Helsinki Declaration.

