## Supplementary Data for "Investigating chromosomal radiosensitivity in inborn errors of immunity: insights from DNA repair disorders and beyond"

**The file includes:**

Supplementary Materials

Supplementary Figures 1-5

Supplementary Tables 1-9

### Supplementary Materials

*Personally identifiable patient information was redacted in accordance with medRxiv requirements.*

#### Protocol for the cytokinesis-block G0 micronucleus assay

For all participants, peripheral whole blood was collected into lithium heparinized tubes and stored at room temperature for a maximum of 24 hours until processing. For each dose point, duplicate cultures were prepared by adding 0.5 ml of whole blood to complete medium consisting of 4 ml RPMI-1640 (Gibco) supplemented with 0.5 ml fetal calf serum (FCS) (Gibco), 50 U/ml penicillin (Gibco), and 50 mg/ml streptomycin (Gibco). Irradiation of blood cultures was performed at the Small Animal Radiation Research Platform (SARRP, Xstrahl, located at Infinity Lab, Ghent University) at room temperature with 0.5 and 1 Gy X-rays (220 kV, 13 mA, 0.15 mm Cu). A square field collimator (100 x 100 mm at 35 cm FSD) was used and the applied dose rate was 3 Gy/min. Physical dosimetry was performed with a pencil ionization chamber (PTW waterproof Farmer chamber (30006)). Sham-irradiated cultures were set up to determine spontaneous MN yields. Phytohemagglutinin (PHA) (Gibco) was added to the blood cultures shortly after irradiation to stimulate lymphocyte proliferation and the cultures were incubated at 37°C, 5% $CO_2$. 23 hours after stimulation, cytokinesis was blocked with 6 mg/ml cytochalasin B (Sigma-Aldrich). Samples were harvested after a total culturing time of 70h and were subsequently hypotonically treated with 0.075 M cold KCl (4°C). The cell suspension was centrifuged, and the remaining cells were fixed with a methanol:acetic acid:Ringer solution (4:1:5). Fixation was repeated twice with methanol:acetic acid solution (4:1) after overnight storage at 4°C. Slides were prepared by dropping cells onto clean slides, followed by staining with 10 mg/ml acridine orange (Sigma-Aldrich). MN were scored manually on a fluorescence microscope by two independent scorers. Per duplicate sample, 500 binucleated (BN) cells were scored, resulting in a total of 1000 BN cells per radiation dose. MN were scored in accordance with internationally accepted scoring criteria.

### Supplementary Figures

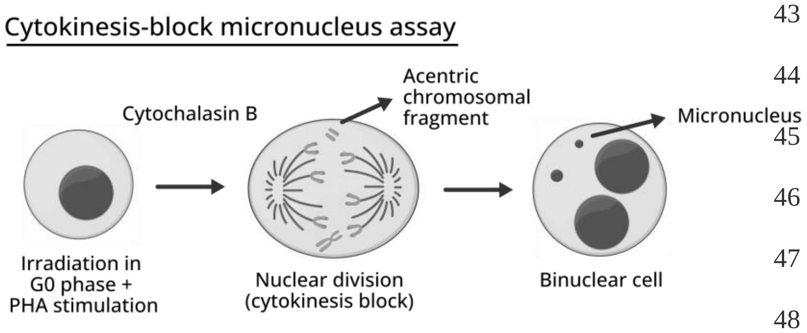


**Fig. S1. Schematic of the G0 cytokinesis-block micronucleus assay (MN) assay.**

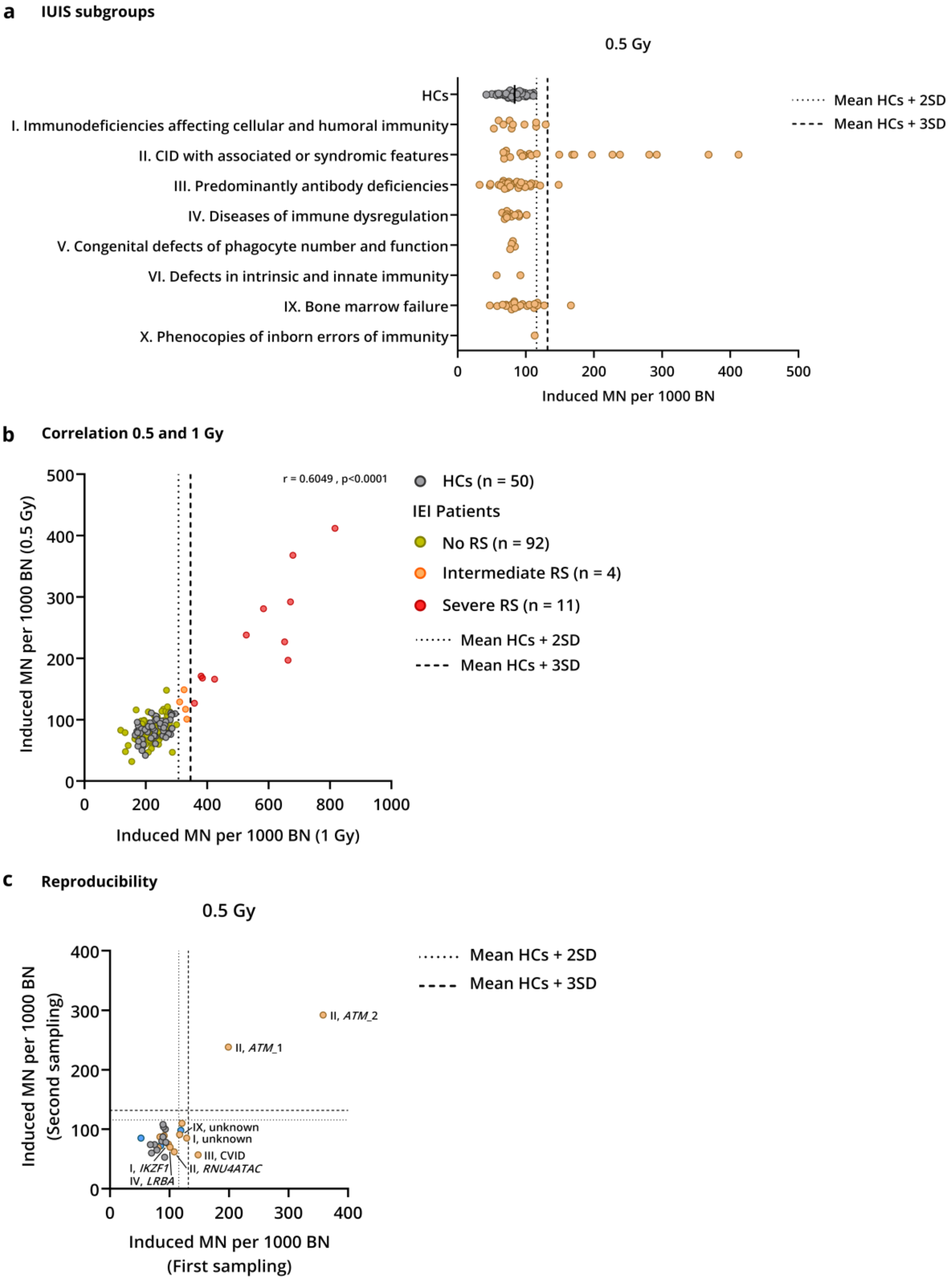


**Fig. S2.**

(**a**) Radiation-induced (0.5 Gy) MN yields are displayed per IUIS subgroup for all patients. (**b**) Comparison of radiation-induced MN yields after 0.5 Gy and 1 Gy exposure. Threshold values were established based on the reference healthy control (HCs) population. Radiation-induced MN yields after 1 Gy were used to determine the chromosomal radiosensitivity (RS) status of the patients. Spearman's rank correlation coefficient (r) between the 0.5 and 1 Gy radiation doses was calculated. Dotted and dashed lines indicate the mean + 2SD and mean + 3SD threshold values, respectively.

**(c)** Reproducibility of the G0 MN assay was assessed by comparing radiation-induced (0.5 Gy) MN yields of the first and second sampling for 11 healthy controls, 3 heterozygous carriers, and 12 patients. Dotted and dashed lines indicate the mean + 2SD and mean + 3SD threshold values, respectively.

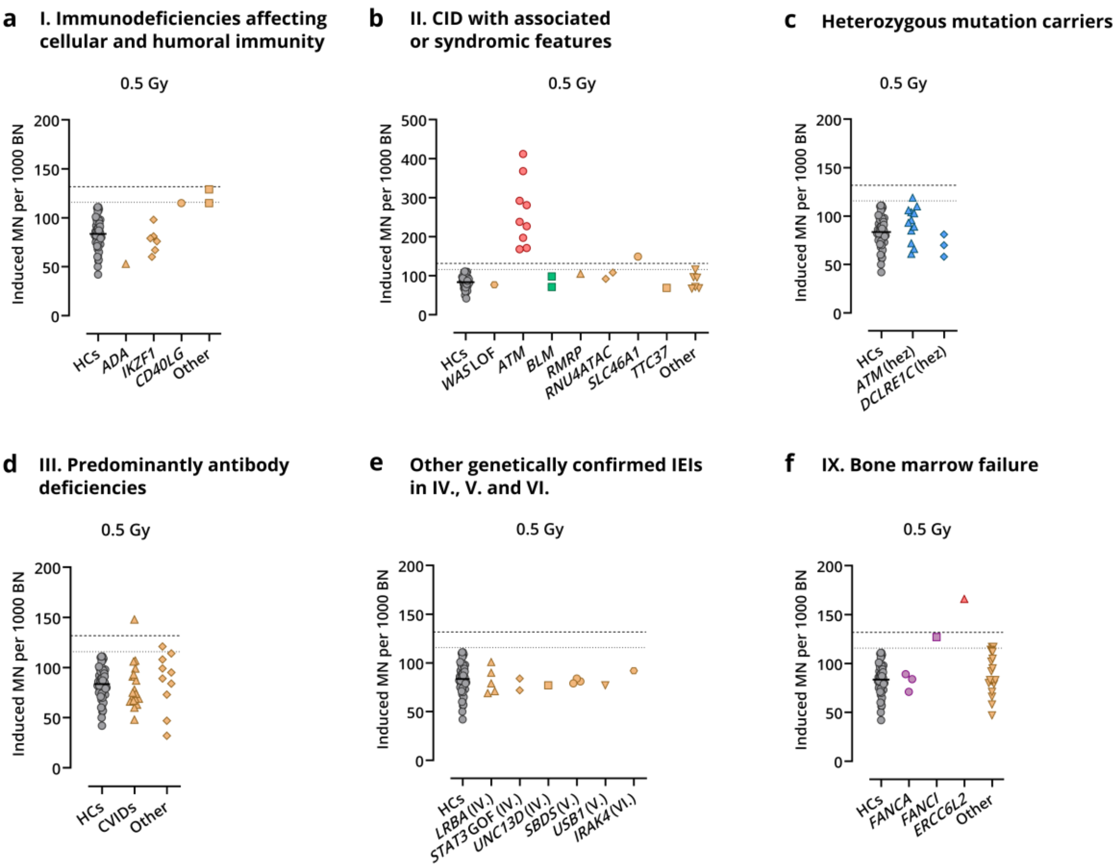


**Fig. S3.**

Radiation-induced MN yields (0.5 Gy) are shown for healthy controls (HCs), heterozygous mutation carriers and patients with a confirmed genetic diagnosis, displayed per IUIS group. **(a)** Patients with a confirmed genetic defect in IUIS group I (immunodeficiencies affecting cellular and humoral immunity) **(b)** Patients with an identified defect in IUIS group II (combined immunodeficiency (CID) with associated or syndromic features). **(c)** Relatives of IEI patients carrying a heterozygous mutation in *ATM* or *DCLRE1C* (Artemis). **(d)** Patients classified in group III (predominantly antibody deficiencies), displayed separately for common variable immunodeficiency (CVID) and non-CVID (other) patients. **(e)** Patients with a confirmed genetic defect in IUIS group IV (diseases of immune dysregulation), group V (congenital defects in phagocyte number and function) or group VI (defects in intrinsic and innate immunity). **(f)** Patients with an identified genetic defect, categorized in IUIS group IX (bone marrow failure). Dotted and dashed lines indicate the mean + 2SD and mean + 3SD threshold values, respectively.

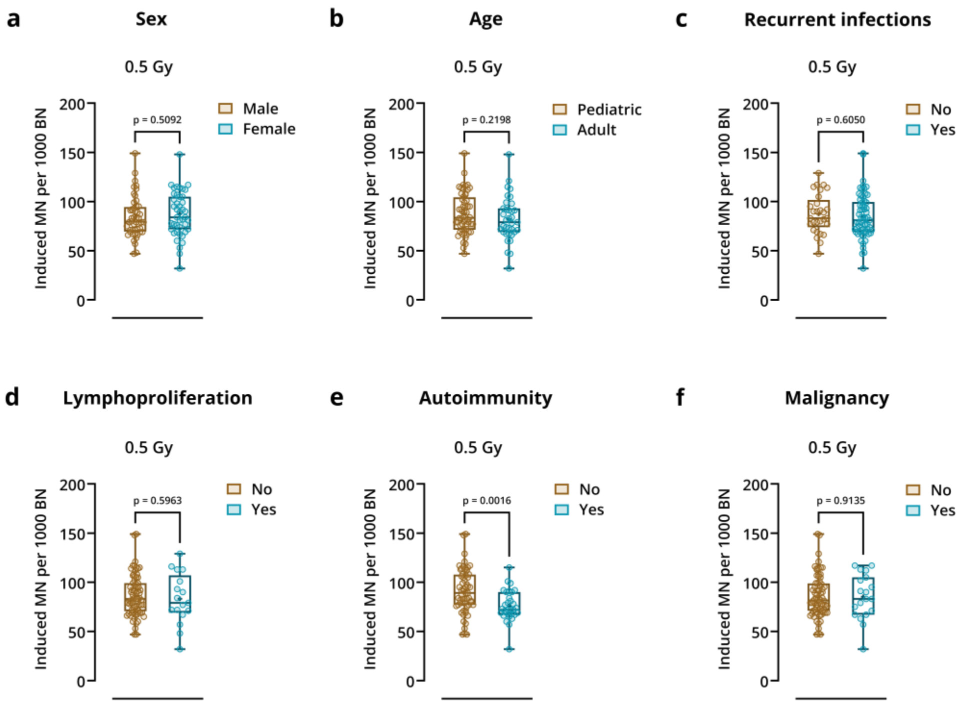


**Fig. S4.**

The association between MN yields and the following clinical features and immunological manifestations were investigated: (**a**) sex, (**b**) age at inclusion, (**c**) infection susceptibility, (**d**) lymphoproliferation, (**e**) autoimmunity, and (**f**) history of malignancy. Radiation-induced MN yields (0.5 Gy) are displayed for each parameter. Boxplots display the median, lower and upper quartiles of the MN yields, whiskers indicate minimum and maximum values. Statistical significance was tested with unpaired t-tests. Patients with a confirmed defect in one of the DNA DSB repair-related genes were excluded for this analysis.

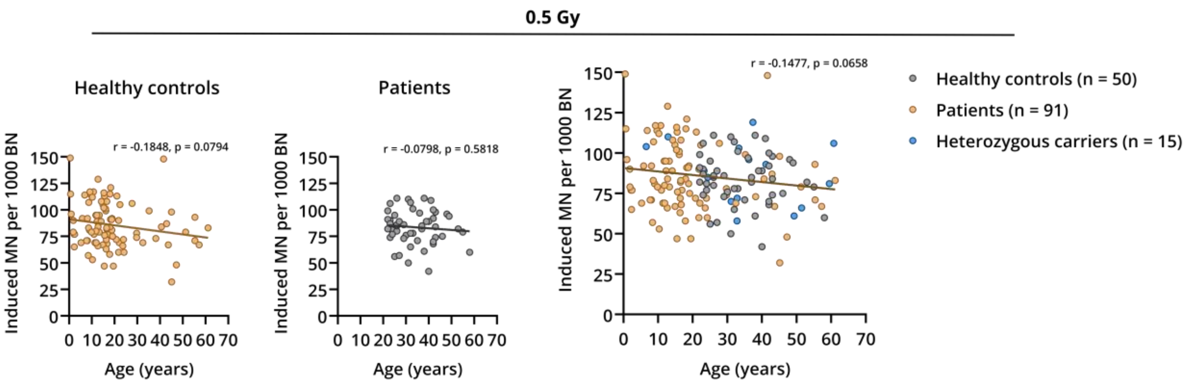


**Fig. S5.**

A correlation analysis was performed between age and radiation-induced MN yields (0.5 Gy), by calculating the Spearman's rank correlation coefficient (r). Both the healthy control and IEI patient cohort were analyzed separately, as well as the entire study population together (healthy controls, patients and heterozygous carriers). Patients with a confirmed defect in one of the DNA DSB repair-related genes were excluded for this analysis.

88 # Supplementary Tables

89 **Table S1. Overview of malignancies reported in the IEI patient cohort.**

| Sex | Genetic IEI diagnosis | Type of malignancy | Current status | Therapy* | Time of RS sampling** |
|---|---|---|---|---|---|
| M | | Hodgkin lymphoma | In remission | Chemotherapy | Pre-diagnosis |
| M | | Non-Hodgkin B cell lymphoma | In remission | Chemotherapy | Pre-therapy |
| M | | Hodgkin lymphoma | In remission | Chemotherapy | Post-therapy |
| M | | Hodgkin lymphoma | In remission | Chemotherapy + Radiotherapy | Post-therapy |
| M | | Non-Hodgkin lymphoma \| Diffuse large B cell lymphoma | In remission | Chemotherapy | Pre-therapy |
| M | | Non-Hodgkin lymphoma \| Diffuse large B cell lymphoma | In remission | Chemotherapy | Post-therapy |
| F | *FANCI* | Secondary acute myeloid leukemia | Deceased | Chemotherapy | Pre-diagnosis |
| F | *LRBA* | Hodgkin lymphoma | In remission | Chemotherapy | Post-therapy |
| F | | Non-Hodgkin lymphoma \| Intracerebral B cell lymphoma | In remission | Radiotherapy | Post-therapy |
| M | | B cell acute lymphoblastic leukemia | In remission | Chemotherapy | Post-therapy |
| M | | Acute myeloid leukemia | In remission | HSCT | Pre-therapy |
| F | *BLM* | Nephroblastoma (Wilms tumor) | In remission | Chemotherapy | Post-therapy |
| F | | Myelodysplastic syndrome - EB2 | In remission | HSCT | Pre-therapy |
| F | | Myelodysplastic syndrome | - | - | - |
| F | | Myelodysplastic syndrome | In remission | HSCT | Pre-therapy |
| M | | Myelodysplastic syndrome | In remission | HSCT | Pre-therapy |
| F | | Myelodysplastic syndrome | In remission | HSCT | Pre-therapy |
| F | | Myelodysplastic syndrome | - | - | - |
| F | | Myelodysplastic syndrome | Deceased | HSCT | Pre-therapy |
| M | | Myelodysplastic syndrome | - | - | - |
| M | | Myelodysplastic syndrome | - | - | - |

90 *M: Male; F: Female; HSCT: Hematopoietic stem cell transplantation; *Only chemotherapy, radiotherapy, or HSCT were recorded; **Only noted when therapy was recorded*

**Table S2. Identified pathogenic variants from IEI patients in group I (Immunodeficiencies affecting cellular and humoral immunity).**

| Gene, Reference sequence | OMIM, inheritance | Patients (n) | IUIS | Nucleotide change | Amino acid change | Consequence | Classification # (ClinVar accession) |
|---|---|---|---|---|---|---|---|
| ***ADA***, NM_000022.4 | 608958, AR | 1 | I.2 | c.[956_960del]; [484T>A] | p.[(Glu319GlyfsTer3)]; [(Ser162Thr)] | Frameshift; Missense | P (VCV000193544.45); LP (Not reported in ClinVar) |
| ***CD40LG***, NM_000074.3 | 308230, XL | 1 | I.2 | c.[761C>T] | p.[(Thr254Met)] | Missense | P (VCV000035814.13) |
| ***IKZF1***, NM_006060.6 | 603023, AD | 6 | I.3 | c.[136del] | p.[(Ser46AlafsTer14)] | Frameshift | P/LP (Not reported in ClinVar) |

*P: Pathogenic; LP: Likely pathogenic; VUS: Variant of uncertain significance*

*# Variant classification noted in ClinVar is shown. For all novel variants not reported in ClinVar, the in-house classification is shown.*

**Table S3. Identified pathogenic variants from IEI patients in group II (Combined immunodeficiency with associated or syndromic features).**

| Gene,<br>Reference<br>sequence | OMIM,<br>inheritance | Patients<br>(n) | Age (yr.),<br>sex | IUIS | Nucleotide change | Amino acid change | Consequence | Classification #<br>(ClinVar accession) |
|---|---|---|---|---|---|---|---|---|
| ***WAS*** LOF,<br>NM_000377.3 | 300392,<br>XL | 1 | >10, M | II.1 | c.[361_365del] | p.[(Asp121ProfsTer46)] | Frameshift | P/LP (Not reported in ClinVar) |
| ***BLM*,**<br>NM_000057.4 | 210900,<br>AR | 1 | 10-19, M | II.2 | c.[3379C>T];<br>[3020-258A>G] | p.[(Gln1127Ter)];<br>[(Met1007fsTer)] | Nonsense;<br>Splice variant | P (VCV000978066.5);<br>P/LP (VCV000978065.6) |
| ***BLM*,**<br>NM_000057.4 | 210900,<br>AR | 1 | >10, M | II.2 | c.[2872G>A];<br>[2872G>A] | p.[(Val958Met)];<br>[(Val958Met)] | Missense | VUS (VCV000650294.19) |
| ***ATM***,<br>NM_000051.4 | 208900,<br>AR | 1 | 10-19, F | II.2 | c.[4490T>G];<br>[8494C>T] | p.[(Leu1497Ter)];<br>[(Arg2832Cys)] | Nonsense;<br>Missense | P/LP (VCV002056154.4);<br>LP (VCV000127459.78) |
| ***ATM***,<br>NM_000051.4 | 208900,<br>AR | 1 | >10, M | II.2 | c.[8494C>T];<br>[1564_1565del] | p.[(Arg2832Cys)];<br>[(Glu522IlefsTer43)] | Missense;<br>Frameshift | LP (VCV000127459.78);<br>P (VCV000127340.74) |
| ***ATM***,<br>NM_000051.4 | 208900,<br>AR | 1 | >10, F | II.2 | c.[1617dup];<br>[5986G>T] | p.[(Cys540MetfsTer26)];<br>[(Glu1996Ter)] | Frameshift;<br>Nonsense | LP (VCV002505532.1);<br>P (VCV000870850.20) |
| ***ATM***,<br>NM_000051.4 | 208900,<br>AR | 1 | >10, M | II.2 | c.[8264_8268del];<br>[8494C>T] | p.[(Tyr2755CysfsTer12)];<br>[(Arg2832Cys)] | Frameshift;<br>Missense | P (VCV000181865.38);<br>LP (VCV000127459.78) |
| ***ATM***,<br>NM_000051.4 | 208900,<br>AR | 1 | >10, F | II.2 | c.[8395_8404del];<br>[5515C>T] | p.[(Phe2799LysfsTer4)];<br>[(Gln1839Ter)] | Frameshift;<br>Nonsense | P/LP (VCV000186242.72);<br>P/LP (VCV000189177.18) |
| ***ATM***,<br>NM_000051.4 | 208900,<br>AR | 1 | 20-29, F | II.2 | c.[7179T>G];<br>[7179T>G] | p.[(Phe2393Leu)];<br>[(Phe2393Leu)] | Missense | VUS* (VCV000407611.10) |
| ***ATM***,<br>NM_000051.4 | 208900,<br>AR | 1 | >10, M | II.2 | c.[1564_1565del];<br>[5511_5512del] | p.[(Glu522IlefsTer43)]:<br>[(Phe1837LeufsTer11)] | Frameshift;<br>Frameshift | P (VCV000127340.74);<br>P/LP (VCV000453583.31) |
| **ATM**,<br>NM_000051.4 | 208900,<br>AR | 1 | 10-19, F | II.2 | c.[8876_8879del];<br>[(2124+1_2125-1)_(2638+1_2639+1)del] | p.[(Asp2959GlyfsTer3)];<br>[(Ile709ValfsTer6)] | Frameshift;<br>Frameshift | P/LP (VCV000189140.37);<br>P (Not reported in ClinVar) |
| ***ATM***,<br>NM_000051.4 | 208900,<br>AR | 1 | 10-19, F | II.2 | c.[3894dup];<br>[8659C>G] | p.[(Ala1299CysfsTer3)];<br>[(His2887Asp)] | Frameshift;<br>Missense | P (VCV000141534.29);<br>VUS (VCV000642280.10) |
| ***RMRP***,<br>NR_003051.3 | 157660,<br>AR | 1 | 10-19, F | II.4 | n.[64C>T];<br>[127C>T] | / | / | P/LP (VCV000189086.15);<br>VUS (VCV000955961.7) |
| ***RNU4ATAC***,<br>NR_023343.1 | 601428,<br>AR | 2 | 10-19, F<br>10-19, M | II.4 | n.[13C>T];<br>[116A>T] | / | / | P/LP/VUS (VCV000218083.30)<br>LP/VCV000812960.3 |
| ***SLC46A1***,<br>NM_080669.6 | 229050,<br>AR | 1 | >10, M | II.6 | c.[616C>T];<br>[649_654dup] | p.[(Gln206Ter)];<br>[(Ala217_Leu218dup)] | Nonsense;<br>In-frame insertion | P (Not reported in ClinVar);<br>VUS (VCV002083625.2) |
| ***TTC37***,<br>NM_014639.4 | 222470,<br>AR | 1 | >10, M | II.9 | c.[643-2A>G];<br>[643-2A>G] | p.[?];<br>[?] | ? | P (Not reported in ClinVar) |

*LOF: Loss-of-function; P: Pathogenic; LP: Likely pathogenic; VUS: Variant of uncertain significance. *G0 MN assay contributed to confirming the pathogenicity of the variant.*

*# Variant classification noted in ClinVar is shown. For all novel variants not reported in ClinVar, the in-house classification is shown.*

**Table S4. Identified pathogenic variants from IEI patients in group III (Predominantly antibody deficiencies).**

| Gene, Reference sequence | OMIM, inheritance | Patients (n) | IUIS | Nucleotide change | Amino acid change | Consequence | Classification # (ClinVar accession) |
|---|---|---|---|---|---|---|---|
| ***NFKB1***, NM_003998.4 | 164011, AD | 1 | III.2 | c.[850C>T] | p.[(Arg284Ter)] | Nonsense | P/LP (VCV000827726.6) |
| ***PIK3CD*** GOF, NM_005026.5 | 615513, AD | 1 | III.2 | c.[3061G>A] | p.[(Glu1021Lys)] | Missense | P (VCV000088675.45) |

*GOF: Gain-of-function; P: Pathogenic; LP: Likely pathogenic*

*# Variant classification noted in ClinVar is shown. For all novel variants not reported in ClinVar, the in-house classification is shown.*

**Table S5. Identified pathogenic variants from IEI patients in group IV (Diseases of immune dysregulation).**

| Gene, Reference sequence | OMIM, inheritance | Patients (n) | IUIS | Nucleotide change | Amino acid change | Consequence | Classification # (ClinVar accession) |
|---|---|---|---|---|---|---|---|
| ***UNC13D***, NM_199242.3 | 608897, AR | 1 | IV.1 | c.[1828_1839del]; [1828_1839del] | p.[(Arg610_Gln613del)]; [(Arg610_Gln613del)] | In-frame deletion | P (VCV000001996.4) |
| ***STAT3*** GOF, NM_139276.3 | 102582, AD | 2 | IV.3 | c.[2147C>T] | p.[(Thr716Met)] | Missense | P (VCV000224848.32) |
| ***LRBA***, NM_001364905.1 | 606453, AR | 2 | IV.3 | c.[6544C>T]; [6544C>T] | p.[(Gln2182Ter)]; [(Gln2182Ter)] | Nonsense | P (Not reported in ClinVar) |
| ***LRBA***, NM_001364905.1 | 606453, AR | 1 | IV.3 | c.[645+22A>C]; [6053A>G] | p.[?]; [(Asp2018Gly)] | ?; Missense | LB/VUS (Not reported in ClinVar); VUS (VCV000951846.9) |
| ***LRBA***, NM_001364905.1 | 606453, AR | 1 | IV.3 | c.[(3825+1_3826-1)_(4339+1_4340-1)del]; [(3825+1_3826-1)_(4339+1_4340-1)del] | p.[(Ile1276Phefs*6)]; [(Ile1276Phefs*6)] | Deletion | P (Not reported in ClinVar) |
| ***LRBA***, NM_001364905.1 | 606453, AR | 1 | IV.3 | c.[3276_3277del]; [3964del] | p.[(Glu1092AspfsTer7)]; [(Leu1322TyrfsTer5)] | Frameshift; Frameshift | P (Not reported in ClinVar); P (Not reported in ClinVar) |

*GOF: Gain-of-function; P: Pathogenic; VUS: Variant of uncertain significance; LB: Likely benign*

*# Variant classification noted in ClinVar is shown. For all novel variants not reported in ClinVar, the in-house classification is shown.*

**Table S6. Identified pathogenic variants from IEI patients in group V (Congenital defects of phagocyte number or function).**

| Gene, Reference sequence | OMIM, inheritance | Patients (n) | IUIS | Nucleotide change | Amino acid change | Consequence | Classification # (ClinVar accession) |
|---|---|---|---|---|---|---|---|
| ***SBDS***, NM_016038.4 | 607444, AR | 1 | V.1 | c.[258+2T>C]; [258+2T>C] | p.[?]; [?] | Splice variant | P/LP (VCV000003196.90) |
| ***SBDS***, NM_016038.4 | 607444, AR | 2 | V.1 | c.[258+2T>C]; [184A>T] | p.[?]; [(Lys62Ter)] | Splice variant; Nonsense | P/LP (VCV000003196.90); P/LP (VCV000449095.18) |
| ***USB1***, NM_024598.4 | 613276, AR | 1 | V.1 | c.[243G>A]; [243G>A] | p.[(Trp81Ter)]; [(Trp81Ter)] | Nonsense | P (VCV000156347.5) |

*P: Pathogenic; LP: Likely pathogenic*

*# Variant classification noted in ClinVar is shown. For all novel variants not reported in ClinVar, the in-house classification is shown.*

**Table S7. Identified pathogenic variants from IEI patients in group VI (Defects in intrinsic and innate immunity).**

| Gene, Reference sequence | OMIM, inheritance | Patients (n) | IUIS | Nucleotide change | Amino acid change | Consequence | Classification # (ClinVar accession) |
|---|---|---|---|---|---|---|---|
| ***IRAK4***, NM_016123.4 | 606883, AR | 1 | VI.7 | c.[79G>C]; [79G>C] | p.[(Asp27His)]; [(Asp27His)] | Missense | VUS (Not reported in ClinVar) |

*VUS: Variant of uncertain significance*

*# Variant classification noted in ClinVar is shown. For all novel variants not reported in ClinVar, the in-house classification is shown.*

**Table S8. Identified pathogenic variants from IEI patients in group IX (Bone marrow failure).**

| Gene, Reference sequence | OMIM, inheritance | Patients (n) | Age (yr.), sex | Nucleotide change | Amino acid change | Consequence | Classification # (ClinVar accession) |
|---|---|---|---|---|---|---|---|
| ***FANCA***, NM_000135.4 | 227650, AR | 1 | 10-19, F | c.[1144C>T];<br>[2505-3C>G] | p.[(Gln382Ter)];<br>[?] | Nonsense;<br>Splice variant | P (VCV001071067.8);<br>VUS (Not reported in ClinVar) |
| ***FANCA,*** NM_000135.4 | 227650, AR | 1 | >10, M | c.[3391A>G];<br>[(2601+1_2602-1)_(2778-1_2779-1)del] | p.[(Thr1131Ala)]:<br>[Phe868_His926del] | Missense;<br>Deletion | P/LP (VCV000237048.50);<br>LP (Not reported in ClinVar) |
| ***FANCA***, NM_000135.4 | 227650, AR | 1 | 10-19, M | c.[2571C>A];<br>[(1470+1_1471-1)_(1626+1_1627-1)del] | p.[(Cys857Ter)];<br>[His492PhefsTer3] | Nonsense;<br>Deletion | P (VCV002445740.1);<br>P (Not reported in ClinVar) |
| ***FANCI***, NM_001376911.1 | 609053, AR | 1 | 40-49, F | c.[3513_3515del];<br>[3931_3941dup] | p.[(Thr1172del)];<br>[(His1314GlnfsTer2)] | In-frame deletion;<br>Frameshift | VUS (Not reported in ClinVar)*;<br>VUS (Not reported in ClinVar)* |
| ***ERCC6L2***, NM_020207.7 | 615667, AR | 1 | >10, F | c.[2367del];<br>[2367del] | p.[(Leu790TyrfsTer62)];<br>[(Leu790TyrfsTer62)] | Frameshift | P (Not reported in ClinVar); |

*P: Pathogenic; LP: Likely pathogenic; VUS: Variant of uncertain significance. *Mitomycin C sensitivity test contributed to confirming the pathogenicity of the variants.*

*# Variant classification noted in ClinVar is shown. For all novel variants not reported in ClinVar, the in-house classification is shown.*

**Table S9. Overview of heterozygous carriers of a (likely) pathogenic *ATM* variant.**

| Age (yr.), gender | Nucleotide change | Amino acid change | Consequence |
|---|---|---|---|
| 20-29, F | c.8494C>T | p.(Arg2832Cys) | Missense |
| 30-39, M | c.1564_1565del | p.(Glu522IlefsTer43) | Frameshift |
| 30-39, F | c.8264_8268del | p.(Tyr2755CysfsTer12) | Frameshift |
| 40-49, M | c.5986G>T | p.(Glu1996Ter) | Nonsense |
| 30-39, F | c.1617dup | p.(Cys540MetfsTer26) | Frameshift |
| 40-49, M | c.8395_8404del | p.(Phe2799LysfsTer4) | Frameshift |
| 30-39, F | c.5515C>T | p.(Gln1839Ter) | Nonsense |
| <10, M | c.1617dup | p.(Cys540MetfsTer26) | Frameshift |
| 10-19, M | c.5986G>T | p.(Glu1996Ter) | Nonsense |
| 20-29, F | c.7179T>G | p.(Phe2393Leu) | Missense |
| 50-59, F | c.7179T>G | p.(Phe2393Leu) | Missense |
| 60-69, F | c.7179T>G | p.(Phe2393Leu) | Missense |

*Reference sequence: NM_000051.4 (ATM)*
